# Assessing the accuracy of a digital symptom checker tool for suggestion of reproductive health conditions: a clinical vignettes study

**DOI:** 10.1101/2023.02.22.23286305

**Authors:** Kimberly Peven, Aidan Wickham, Octavia Wilks, Yusuf C. Kaplan, Andrei Marhol, Saddif Ahmed, Ryan Bamford, Carley Prentice, Andras Meczner, Matthew Fenech, Stephen Gilbert, Anna Klepchukova, Sonia Ponzo

**Affiliations:** Flo Health UK Limited, London, United Kingdom; Your.MD Limited, London, United Kingdom; Una Health GmbH, Hamburg, Germany; Else Kröner Fresenius Center for Digital Health, Technische Universität Dresden, Dresden, Germany; University College London, Institute of Health Informatics

## Abstract

**Background:** Reproductive health conditions such as endometriosis, uterine fibroids and polycystic ovary syndrome affect a large proportion of women and people who menstruate worldwide. Prevalence estimates for these conditions range from 5-40% of women of reproductive age. Long diagnostic delays, up to 12 years, are common and contribute to health complications and increased healthcare costs. Symptom checker apps provide users with information and tools to better understand their symptoms and thus have the potential to reduce the time to diagnosis for reproductive health conditions.

**Objective:** This study aims to evaluate the accuracy of three symptom checkers developed by Flo Health assessing symptoms of endometriosis, uterine fibroids and polycystic ovary syndrome (PCOS) against current medical guidelines.

**Methods:** Independent general practitioners were recruited to create clinical case vignettes of simulated users with and without the conditions of interest. Vignettes were reviewed, modified and approved by separate general practitioners. A further independent panel of general practitioners reviewed the cases and designated a final classification. Vignettes were entered into the symptom checkers and the outcomes were compared with the final classification from the panel using accuracy metrics including percent agreement, sensitivity and specificity.

**Results:** A total of 24 cases were created per condition. Overall, exact matches between the vignette classification and the symptom checker outcome was 83.3% for endometriosis and uterine fibroids, and 87.5% for PCOS. While sensitivity was high for all conditions (>81%) and very high (100%) for PCOS, specificity was >81% for endometriosis and uterine fibroids and 75% for PCOS.

**Conclusion:** The single condition symptom checkers have high levels of accuracy for endometriosis, uterine fibroids and PCOS. Given long delays in diagnosis for many reproductive health conditions, which lead to increased medical costs and potential health complications for individuals and healthcare providers, innovative health apps and symptom checkers hold the potential to improve care pathways.

## Background

Millions of women and people who menstruate worldwide are affected by reproductive health conditions. Endometriosis, symptomatic uterine fibroids, polycystic ovary syndrome (PCOS) are among the most common with prevalences estimated at 10-15%, 20-40%, and 5-20%, respectively ^1–12^. Endometriosis is a condition where endometrial tissue is found outside of the uterus ^13^. Uterine fibroids are benign uterine tumours, which can cause a variety of debilitating symptoms, such as heavy menstrual bleeding, pain, bladder and/or bowel dysfunction ^12,14^. Both endometriosis and uterine fibroids severely affect quality of life, everyday functioning and workplace productivity ^15–19^. Further, both conditions have been associated with fertility issues ^20^. PCOS is a complex endocrine disorder characterised by a variety of symptoms of differing severity and without a certain aetiology ^21^. Infertility and type 2 diabetes are common sequelae, as are cardiovascular and psychiatric conditions (e.g. hypertension, depression, anxiety)^22^.

Long diagnostic delays are common, with patients reporting receiving a diagnosis between 2 and 12 years from the onset of symptoms ^23–28^. A contributing factor to diagnostic delays is low reproductive health literacy. Affected persons may believe symptoms are normal or hereditary, thus delaying seeking medical input until symptoms worsen ^29^. Controversy over diagnostic criteria may further complicate or delay final diagnosis ^12,30–32^. In addition to risks for developing complications with fertility or psychiatric conditions, ^33–36^ long diagnostic delays are associated with increased healthcare utilisation and costs ^37^. Endometriosis costs an average of $27,855 per patient annually in the US alone^8^, whilst overall yearly expenditure for uterine fibroids is estimated to be $34.4 billion ^19^. Further, patients with long diagnostic delays for endometriosis have 60% higher mean all-cause costs compared to those with short delays ^37^. Similarly, the economic costs of PCOS on individuals and healthcare systems is estimated to be $8 billion per year ^8,19^.

As diagnostic costs represent a small proportion of the total economic burden of disease, particularly in light of long diagnostic delays, access to simpler screening processes may be a cost-effective strategy ^38^. Innovations in health tech and mobile applications (apps) have the potential to bridge this gap. Worldwide, there are more than 6 billion smartphone subscribers ^39^ and more than 350,000 health-related mobile apps ^40^. As such, people increasingly turn to the internet for health information ^41–43^ and demand exists for health screening mobile apps to assist with condition diagnosis (e.g. check user symptoms against common condition symptoms) ^44–46^.

Despite the widespread availability and advantages of symptom checker apps, there remains a knowledge gap on the accuracy of many of these tools ^51^. Researchers, clinicians and patient groups are increasingly demanding more rigorous validation and evaluation of digital health solutions, with scientists highlighting the need for evidence generation ^47–50^. Case vignette studies represent an established methodology for the evaluation of online symptom checkers. In such studies, relevant fictitious patient cases are assessed by the symptom checker under investigation and the output is compared to that of a human expert assessing the same case ^51^. Of the available symptom checkers, some do not provide clear information on their authors, information sources, or evaluation and testing, and reported accuracy metrics vary greatly ^51^. A recent review of online symptom checkers found diagnostic accuracy of the primary diagnosis varied from 19-38% and triage accuracy ranged from 49-90% ^52^. Even though information on their development and validation is limited and its reliability in question ^45,51^, trust in symptom checker apps is high among laypersons ^53^.

The aim of the current study was to determine the accuracy of three symptom checkers assessing symptoms of endometriosis, uterine fibroids and PCOS against current medical guidelines. To this end, we devised a case vignette study whereby fictional patient cases were assessed for symptoms of the above mentioned conditions by both symptom checkers and medical practitioners.

## Methods

### Flo app and symptom checker development

Flo ^54^(by Flo Health UK Limited) is a women’s health and wellbeing mobile app and period-tracker with over 50 million monthly active users. Flo allows users to track their symptoms throughout their menstrual cycle (e.g. cramps, menstrual flow, mood) or pregnancy and postpartum (e.g. lochia), as well as general health information like contraceptive use, ovulation or pregnancy test results, water intake, and sleep. Additionally, the app offers personalised, evidence-based and expert-reviewed content via an in-app library. Further, digital health assistants (“chatbots”) provide users with information about a range of conditions.

Flo has developed three single-condition symptom checker “chatbots” to assess symptoms of reproductive health conditions (endometriosis, uterine fibroids, PCOS). The symptom checkers (not yet publicly available) use symptom information gained through conversation-like question and answers, as well as symptom or menstrual cycle information previously entered into the app. Users with acute presentations are provided with a list of red flag symptoms (e.g. nausea with vomiting, fever, vaginal bleeding not related to the period) at the beginning of the conversation, and are advised to discontinue the conversation with the symptom checker and seek urgent medical advice if their presence is confirmed by the user. After the conversation, the symptom checker gives the user one of two possible outcomes: 1) A strong match for the condition - *“You’re experiencing several symptoms typically associated with [condition]”* or 2) Weak or no match for the condition - *“While you may be experiencing some symptoms of [condition], your combination of symptoms does not strongly indicate it”*. An informative summary is available for the user which reiterates which of the user’s symptoms match the presentation of a particular condition as described in the relevant clinical guideline(s). This summary can then be used by the user to facilitate any subsequent conversations with their healthcare provider. The symptom checker is not intended as a diagnostic tool, does not provide medical advice, and users are advised to seek medical input to further investigate any concerns they have.

To ensure medical accuracy and safety during development of symptom checkers, Flo uses a combination of an in-house medical team and external doctors specialising in the conditions of interest. The medical team builds the chat sequences considering the most relevant signs and symptoms based on the latest medical guidelines and evidence. The chat sequence is medically tested, reviewed, and adjusted in an iterative product development process.

### Vignette testing

Clinical case vignettes were created, reviewed, approved, classified, and entered into the symptom checkers by independent general practitioners (GPs) recruited specifically for this study. All GPs were UK-based with an average of 12 years of clinical experience and were not previously affiliated with Flo. All GPs were remunerated for their time.

#### Vignette creation, review and approval

Five external GPs were recruited to independently create clinical case vignettes of simulated users (Figure 1, step one). These simulated users would be presenting for the first time, without any history of diagnosis or treatment for one of the three conditions of interest, namely endometriosis, uterine fibroids, or PCOS. Cases were derived from the GPs’ clinical experience and the literature. The GPs completed a template (see Supplementary Materials, Appendix one) for each vignette which contained information on the user’s background, history of presenting condition, medical, surgical, and family history, as well as details on their menstrual cycle and other symptoms. The GPs were instructed to create a set number of cases for each of the three conditions, for each of three possible outcomes to ensure a spread of severity and condition types: A) *“You’re experiencing specific signs and symptoms commonly associated with [condition]”*, B) *“Although you’re experiencing some of the potential signs and symptoms of [condition], they are not specific enough to indicate it strongly*.*”*, C) *“You’re not experiencing any of the signs and symptoms commonly associated with [condition]*.*”*. GPs were instructed that “A” cases are those for which the user has specific features of the condition and this differential diagnosis is the most likely cause of their symptoms. For “B” and “C” cases, these are not considered to have the condition. GPs were instructed that “B” cases represent users who show either too few or only some specific fındings, and a clinician would not think of this condition as the most likely cause for these symptoms. “C” cases represent users who show either too few or non-specific symptoms and there would be other differential diagnoses which are more likely to be the cause of the symptoms. Condition-negative cases had other diagnoses such as urinary tract infection, thrush, pregnancy, functional constipation.

**Figure 1.**
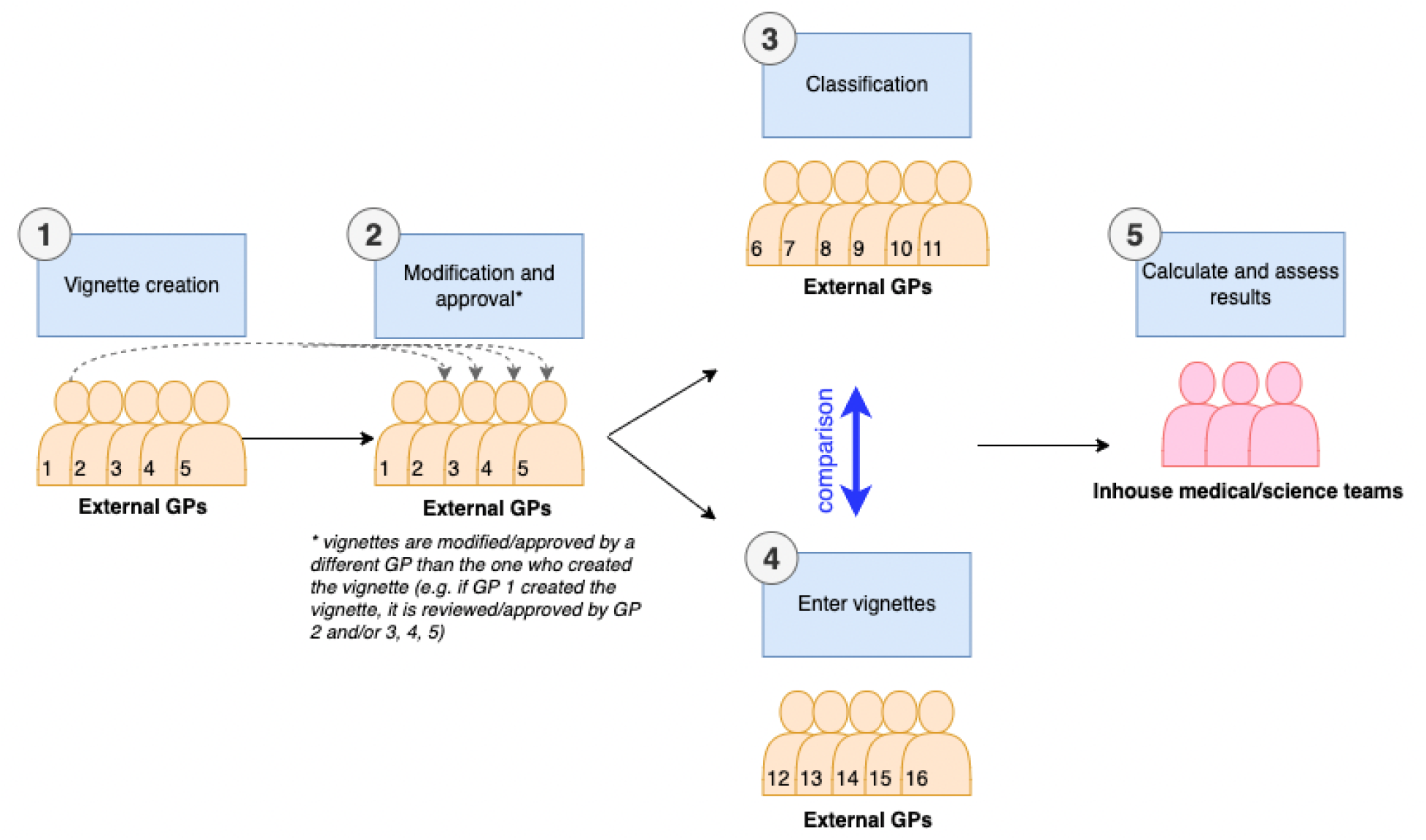
Vignette study procedure including 1) independent vignette creation by five external GPs, 2) review, modification, and approval of vignettes by a second GP and third where required, 3) independent vignette classification by three external GPs not involved in other stages, 4) entry of vignettes into symptom checkers by external GPs not involved in other stages, and 5) analysis of results

Each vignette was reviewed by a second GP (Figure 1, step two) who could either approve the vignette as-is, or suggest changes to clarify the case. If changes were suggested, the case would then be reviewed, edited, and approved by a third GP who would finalise the case. Twenty-four cases were created for each condition, in line with other single-condition or single-system symptom checker evaluations ^55–58^.

#### Independent classification of vignettes

To avoid bias from the case creator setting the final classification, an additional independent panel was recruited to classify the vignettes. After vignette approval (Figure 1, stage 2), the type of case (A, B, or C above) was removed from the vignette template, as were any notes about the diagnosis the creator had in mind when creating the vignette. Six additional external GPs (not involved in step one and two) classified the vignettes (Figure 1, step three). The classifying GPs received a random selection of vignettes, each designated as either an endometriosis vignette, uterine fibroid vignette, or PCOS vignette. For each vignette, the GPs reviewed the case and designated the most likely outcome for the specified condition (endometriosis, uterine fibroids, or PCOS) matching the symptom checker wording: 1) A strong match for the condition - *“You’re experiencing several symptoms typically associated with [condition]”* or 2) Weak or no match for the condition - *“While you may be experiencing some symptoms of [condition], your combination of symptoms does not strongly indicate it”*. Each vignette was reviewed independently by three GPs. The majority view (at least two out of three) was taken as the “true value” for the vignette. While the vignettes were created with three levels of categorisation for each condition, the classifying GPs were not aware of these levels and were asked to make a binary classification for each vignette.

#### Vignette entry

An additional set of five external GPs (not involved in the other steps) were recruited to enter the vignette cases into a prototype of the symptom checkers (Figure 1, step four). At this stage the GPs were blinded to the condition assigned to the vignette, the classification, and the condition the symptom checker was assessing. If the symptom checker asked a question that was not contained in the vignette, GPs were instructed to follow a step-by-step protocol to determine the appropriate answer. First, if the information requested by the symptom checker was specified in the vignette template (e.g. the vignette template specifies pain symptoms should include details on radiation of pain, if applicable) but the information was not included by the creator (e.g. pain was listed but radiation of pain was not mentioned), it could be assumed to not apply and a negative response should be selected. If the information was not part of the template, a neutral response (e.g. “I don’t know”, “I don’t want to answer this question”) should be selected. If no neutral response was available, a negative response should be selected. If no negative response was available, the answer most within normal limits should be selected (e.g. the inputting GP would select a period length of 2-7 days, as opposed to a period length of 1 day or less, or a period length of 8 days or more).

### Analysis

The final classification set by the independent GP classifiers (Figure 1, step three) was compared with the outcome of the symptom checker as tested in Figure 1, step four. Outcomes were arranged in two-way tables as shown in Table 1. Accuracy statistics were calculated using standard formulas as described in Supplementary Materials, Appendix two.

**Table 1:**
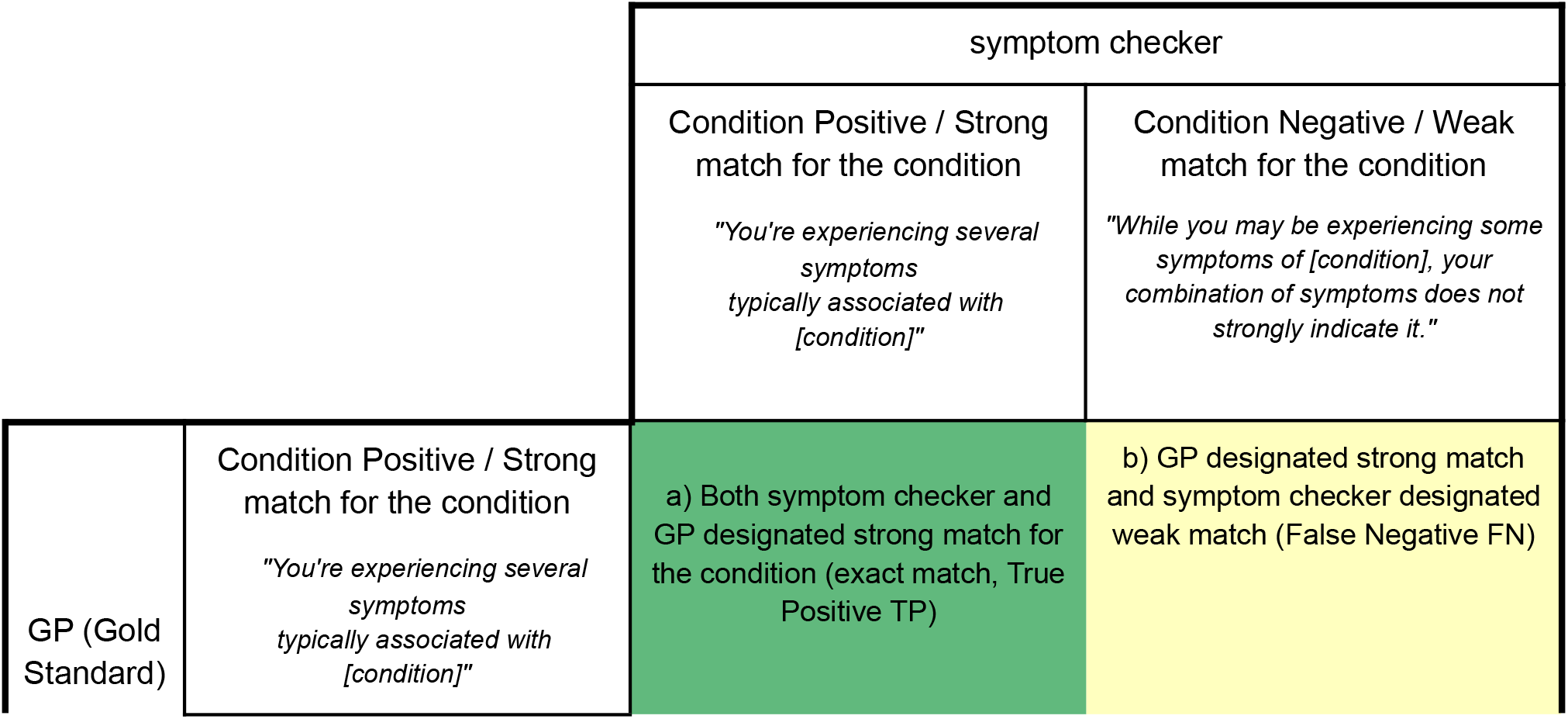
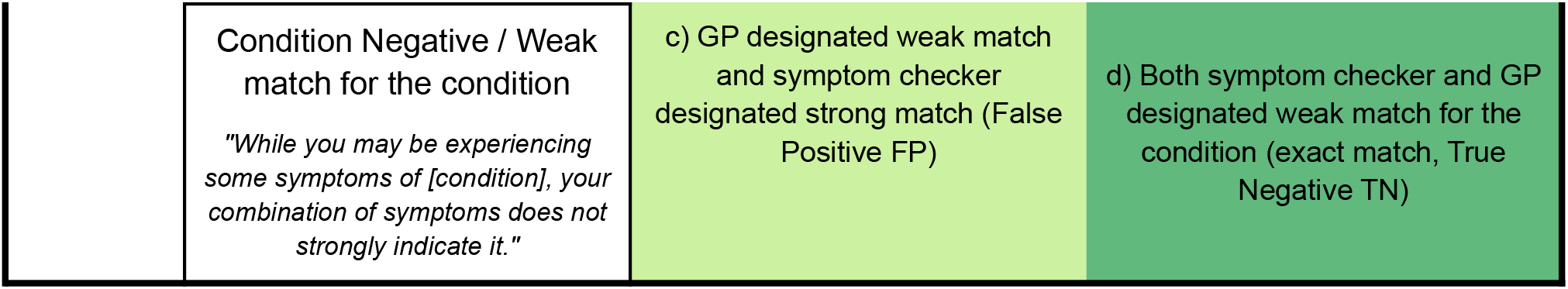
Two-way validation table

## Results

### Vignette cases

Out of the total of 24 cases that were created per condition (Table 2) 11-13 cases were classified as a strong match for the condition and 11-13 cases were classified as a weak match for the condition after final classification by a panel (shown in Figure 1, step 3).

**Table 2 (A-C):** Two-way validation tables by condition

A) Endometriosis
|  |  | Endometriosis symptom checker |  |  |
| --- | --- | --- | --- | --- |
|  |  | Condition Positive / Strong match for the condition | Condition Negative / Weak match for the condition | Total |
| GP (Gold Standard) | Condition Positive / Strong match for the condition | 9 | 2 | 11 |
|  | Condition Negative / Weak match for the condition | 2 | 11 | 13 |
|  | Total | 11 | 13 | 24 |

B) Uterine fibroids
|  |  | Uterine fibroids symptom checker |  |  |
| --- | --- | --- | --- | --- |
|  |  | Condition Positive / Strong match for the condition | Condition Negative / Weak match for the condition | Total |
| GP<br>(Gold<br>Stand<br>ard) | Condition Positive /<br>Strong match for the<br>condition | 11 | 2 | 13 |
|  | Condition Negative /<br>Weak match for the<br>condition | 2 | 9 | 11 |
|  | Total | 13 | 11 | 24 |

C) PCOS
|  |  | PCOS symptom checker |  |  |
| --- | --- | --- | --- | --- |
|  |  | Condition Positive /<br>Strong match for the<br>condition | Condition Negative /<br>Weak match for the<br>condition | Total |
| GP<br>(Gold<br>Stand<br>ard) | Condition Positive /<br>Strong match for the<br>condition | 12 | 0 | 12 |
|  | Condition Negative /<br>Weak match for the<br>condition | 3 | 9 | 12 |
|  | Total | 15 | 9 | 24 |

### Accuracy

Overall, exact matches (percent agreement) between the vignette classification and the symptom checker outcome ranged from 83.3% for endometriosis and uterine fibroids to 87.5% for PCOS (Figure 2, Table 3). While there were no false negative outcomes for PCOS, 8.3% of all cases were falsely identified by the relevant symptom checker as negative for endometriosis and uterine fibroids. False positive outcomes ranged from 8.3% for endometriosis and uterine fibroids to 12.5% of all cases for PCOS.

**Table 3:** Accuracy metrics for Endometriosis, Fibroids and PCOS

| Condition | Number (n) | Agreement (%) | Sensitivity (%) | Specificity (%) | PPV (%) | NPV (%) |
| --- | --- | --- | --- | --- | --- | --- |
| Endometrio<br>sis | 24 | 83.3 | 81.8 | 84.6 | 81.8 | 84.6 |
| Fibroids | 24 | 83.3 | 84.6 | 81.8 | 84.6 | 81.8 |
| PCOS | 24 | 87.5 | 100 | 75 | 80 | 100 |

**Figure 2.**
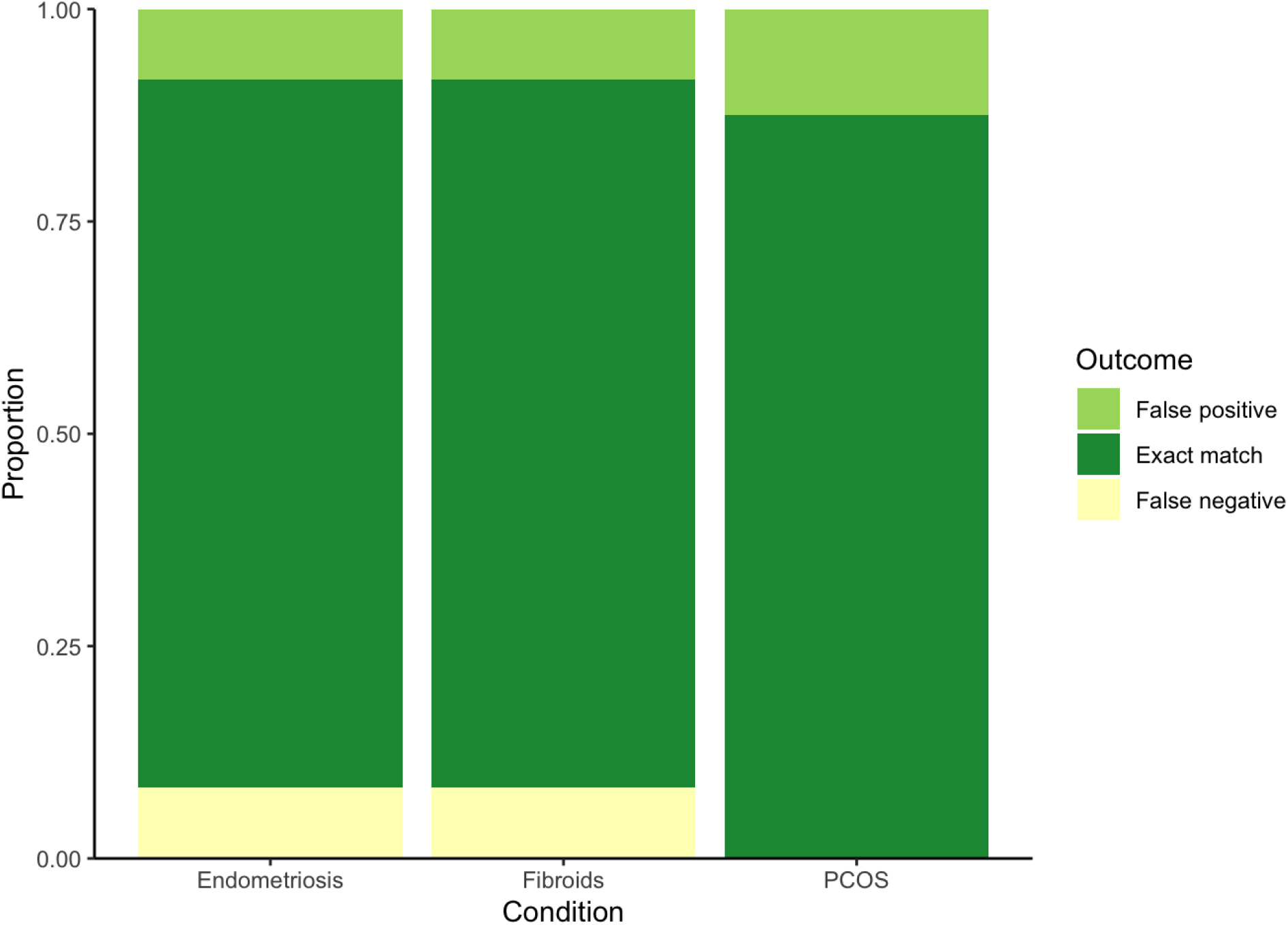
Overall symptom checker performance showing the proportion of false positive outcomes, exact match outcomes, and false negative outcomes by condition.

While sensitivity was very high (100%) for PCOS (Table 3), specificity was high for all three conditions (>81%). Positive predictive value ranged from 80.0% for PCOS to 84.6% for uterine fibroids while negative predictive value ranged from 81.8% for uterine fibroids to 100% for PCOS.

## Discussion

### Summary

In this case vignette study, we assessed the accuracy of three single-condition symptom checkers for three reproductive health conditions (endometriosis, fibroids, PCOS). We found the designation given to case vignettes by the symptom checkers had high levels of accuracy (83.3-87.5%), sensitivity (81.8-100.0%), and specificity (75.0-84.6%) when compared to gold standard GP designation.

### Comparison with prior work

This high accuracy of identification of reproductive health conditions is particularly important as high rates of diagnostic error are reported by patients. A study of patients with self-reported surgically confirmed endometriosis found that 75.2% of patients reported being misdiagnosed with another physical health and/or mental health problem by their health care professional ^59^. A similar study of patients with diagnosed PCOS found that 33.6% of women reported >2 years time to diagnosis and 47.1% visited ≥3 health professionals before a diagnosis was established, and 64.8% were dissatisfied with the diagnostic process ^60^.

Other vignette studies of multi-condition symptom checkers have shown mixed results for accuracy. A study by Gilbert et al ^61^ comparing urgency advice (i.e. triage) from 7 multi-condition symptom checker apps and 7 GPs to gold standard vignettes found the condition suggested first matched the gold standard (i.e. M1 accuracy) for 71% of GPs and 26% of apps; when broadening to the condition suggested in the top-five (i.e. M5 accuracy), GP accuracy rose to 83% and apps to 41%. Another study by Schmieding et al ^62^ comparing 22 symptom checkers using 45 vignettes found M1 accuracy of 46% and M10 accuracy was 71%.

The multi-condition symptom checkers described above were studied with vignettes designed to cover common and less-common conditions seen in primary care practice affecting all body systems and including a range of urgency levels. Further, the symptom checkers evaluated by Gilbert et al and Schmieding et al are designed to detect a wide range of conditions for a general population. In contrast, our study evaluated single-condition symptom checkers using vignettes specifically designed to represent presentations with specific symptoms of the condition (strong match/condition positive) and presentations with symptoms not specific to the condition (weak match/condition negative). This symptom checker design difference may explain variation in accuracy found between our symptom checkers (single-condition) and other studied symptom checkers (multi-condition).

Evaluations of single-condition symptom checkers include a study of 12 web-based symptom checkers for COVID-19 ^63^ and a study of an app-based symptom checker for PCOS ^56^. COVID-19 symptom checkers ranged widely in both sensitivity (14-94) and specificity (29-100), with only four symptom checkers having both sensitivity and specificity above 50% and two with both sensitivity and specificity above 75%. Sensitivity and specificity in our symptom checkers was between 75-100%. The PCOS symptom checker evaluated by Rodriguez et al ^56^ reported 12-25% false-positive cases and no false-negative out of 8 cases tested. Our PCOS symptom checker had no false-negatives and three false-positive cases (12.5%) out of 24 cases tested.

With the exception of COVID-19, which has a symptomatology and overall presentation that differs greatly from the reproductive health disorders assessed in the current study, digital or app-based symptom checkers for a single condition are uncommon. Symptom-based or patient-completed questionnaires or screening tools do exist, including for common reproductive health conditions such as endometriosis or PCOS. A patient self-assessment tool for endometriosis with 21 questions found sensitivity of 76% and specificity of 72% ^64^. Our endometriosis symptom checker had a similar but slightly higher sensitivity (81.8%) and sensitivity (84.6%). A four-item questionnaire for use in diagnosis of PCOS among women with a primary complaint of infertility had 77% sensitivity and 94% specificity ^65^. Our PCOS symptom checker had higher sensitivity (100%) and lower specificity (75%), prioritising identification of cases. It should be noted, however, that our symptom checker is designed to be for a broader population than the four-item clinical tool, including those who are not trying to get pregnant or experiencing fertility issues. Questionnaires such as these have some limitations. They may not be available to the public and additionally may be subject to more user error (e.g. question skipping). App-based symptom checkers, on the other hand, can use historical data from users such as menstrual regularity to improve accuracy of user answers. Additionally, users cannot accidentally skip questions, and the app will provide a detailed summary of results and recommendations.

The possible applications of symptom-checkers and health apps are far-reaching and could have benefits at the individual user-level, healthcare professional level, and macro/health system level ^57,66^. Especially for many reproductive health conditions where time to diagnosis is currently long and contributes to high healthcare costs, ^23,29,60,67^ an earlier diagnosis can lead to early treatment and thus decrease complications from untreated conditions and decrease healthcare costs of treating more advanced disease ^33,34,37^. Additionally, menstrual cycle details such as cycle length, period length, or flow can be important information for healthcare providers when making a diagnosis. Health apps can help track cycle details over time and use these details when determining risk for conditions as well as in summary information for users to share with their health care providers. As people with symptoms such as heavy bleeding or menstrual pain may believe these are normal or hereditary ^29^, personalised assessment of symptoms and encouragement to seek further evaluation from a medical professional where appropriate may improve an individual’s understanding of their symptoms and health status and decrease time to diagnosis. By using a combination of tracked cycle details, symptoms experienced, and medical/family history, symptom checkers could optimise pathways to diagnosis. Particularly where mobile apps with symptom tracking can identify users with risk factors for certain conditions, educating users about their symptoms may encourage conversation with their medical providers. This may be most relevant in populations with low health literacy who may think the symptoms are normal, or who may hesitate to seek care from medical professionals ^29,68^. This is particularly important in minority communities and economically deprived areas where time to diagnosis can be longer and participation in health screening is lower ^68^. Further, some ethnic minority groups have higher rates of using smartphones for health information ^69^ Additionally, health apps may improve patient-provider communication as users can share results and symptom patterns with their care provider (For example, the Flo app provides a “health report” where you can download a summary of symptoms over a period of time, average cycle length, and other details to share with a health care provider).

Other studies have shown variation between groups of GPs reviewing vignettes ^63^. Each vignette in our study was reviewed independently by three different GP classifiers. In 80.5% of cases, all three GPs agreed with each other, while in 19.4% of cases, one of the GPs had a different opinion. This disagreement between GPs and some differences with the symptom checker results are to be expected, particularly when using symptom-based assessment for reproductive health conditions that can be complicated to diagnose, have overlapping symptomatology with other system conditions such as gastrointestinal and urinary conditions, and are often dismissed or considered to be “normal” variations in the menstrual cycle by some. These conditions have notoriously prolonged time to diagnosis ^23–26^ and require investigations including imaging. Further, sensitivity of different testing methods can vary. For example, physical examination for deep infiltrating endometriosis can have poor accuracy and requires imaging ^71^.

### Strengths and limitations

Strengths of this study include the use of different groups of independent, external GPs unfamiliar with the symptom checkers to create, enter, and classify case vignettes for symptom checker testing. Additionally, vignettes were created with a wide range of symptomatology to ensure inclusion of borderline presentations as these are notoriously difficult to assess, even for doctors, although they represent a frequent reality as people do not often fit neatly into textbook case presentations. Further, vignette cases were each reviewed by an independent, experienced GP and classified by a separate panel viewing the vignettes for the first time. We created 72 vignette cases total, 24 for each of our three conditions. The number of vignettes needed to evaluate symptom checkers is not well defined ^72^. Other vignette symptom checker evaluations have used between 3-400 cases for testing with single-condition or single-system evaluations (e.g. mental health, ophthalmology, PCOS) using fewer cases and multi-condition evaluations using larger numbers of cases ^55–57,73,74^. Amongst the 400 vignettes published by Hammoud et al ^74^, any single condition is only represented by at most five cases. Limitations, however, should be noted. Vignette studies rely on clinical opinion of a small number of GPs. An audit study of clinical vignette benchmarking has shown significant variation between groups of GPs considering clinical vignettes ^70^. To decrease bias from difference in clinical opinion, all cases were blindly reviewed by three GPs, a third involved in cases of disagreement. We found agreement between all three GPs in 80.5% of our cases. Vignettes also rely on classical presentation of conditions which may present differently in real life. Additionally, although we recognise that patients do not usually present to primary care practitioner with a pre-specified suspected diagnosis, and that therefore this aspect of the study design does not reflect usual medical practice, these chatbots are not meant to replace the interaction with primary care providers but rather to allow users to review their symptoms in advance of seeing a healthcare professional. While we found 100% sensitivity for our PCOS symptom checker, it is likely with a larger sample size and real-life cases, this level of perfect sensitivity will not be maintained.

Other changes in accuracy statistics are likely to be seen in real-world use. Further, as real-world users may interpret their symptoms and the questions differently than doctors, future studies including the general population should be carried out. Evaluation of symptom checkers and digital health tools should follow multistage processes with increasing exposure to real environments exploring not only effectiveness but also usability and exploring balance between probability of disease and risk of missing a diagnosis ^75^.

## Conclusions

In conclusion, we found high levels of accuracy for single-condition symptom checkers for three reproductive health conditions. Given long delays in diagnosis for many reproductive health conditions, which lead to increased medical costs and potential health complications, innovative health apps and symptom checkers hold the potential to improve care pathways.

## Data Availability

All data produced in the present study are available upon reasonable request to the authors

## Acknowledgements

This study was funded by Flo Health UK Limited

## Supplementary material

**Appendix 1.**
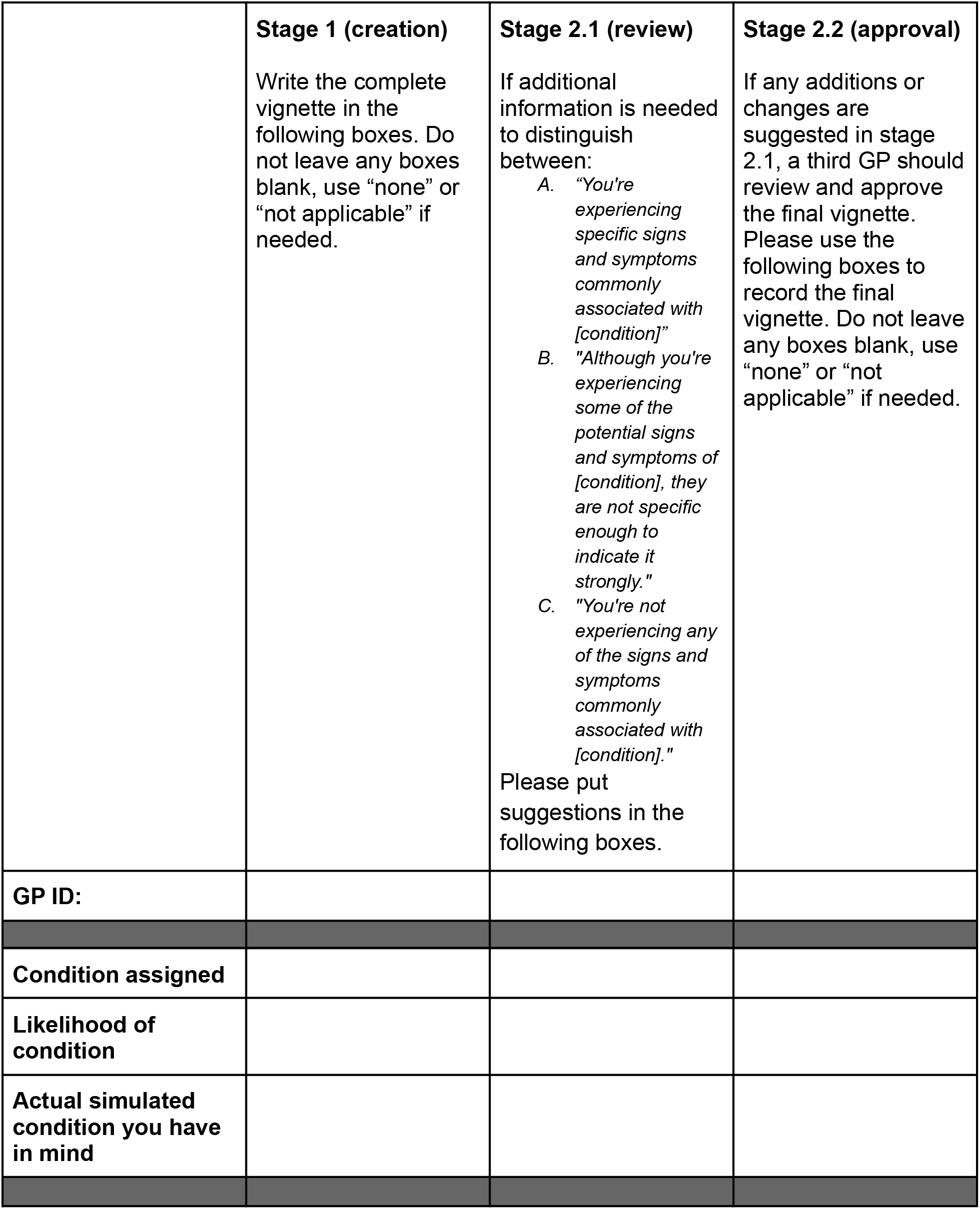
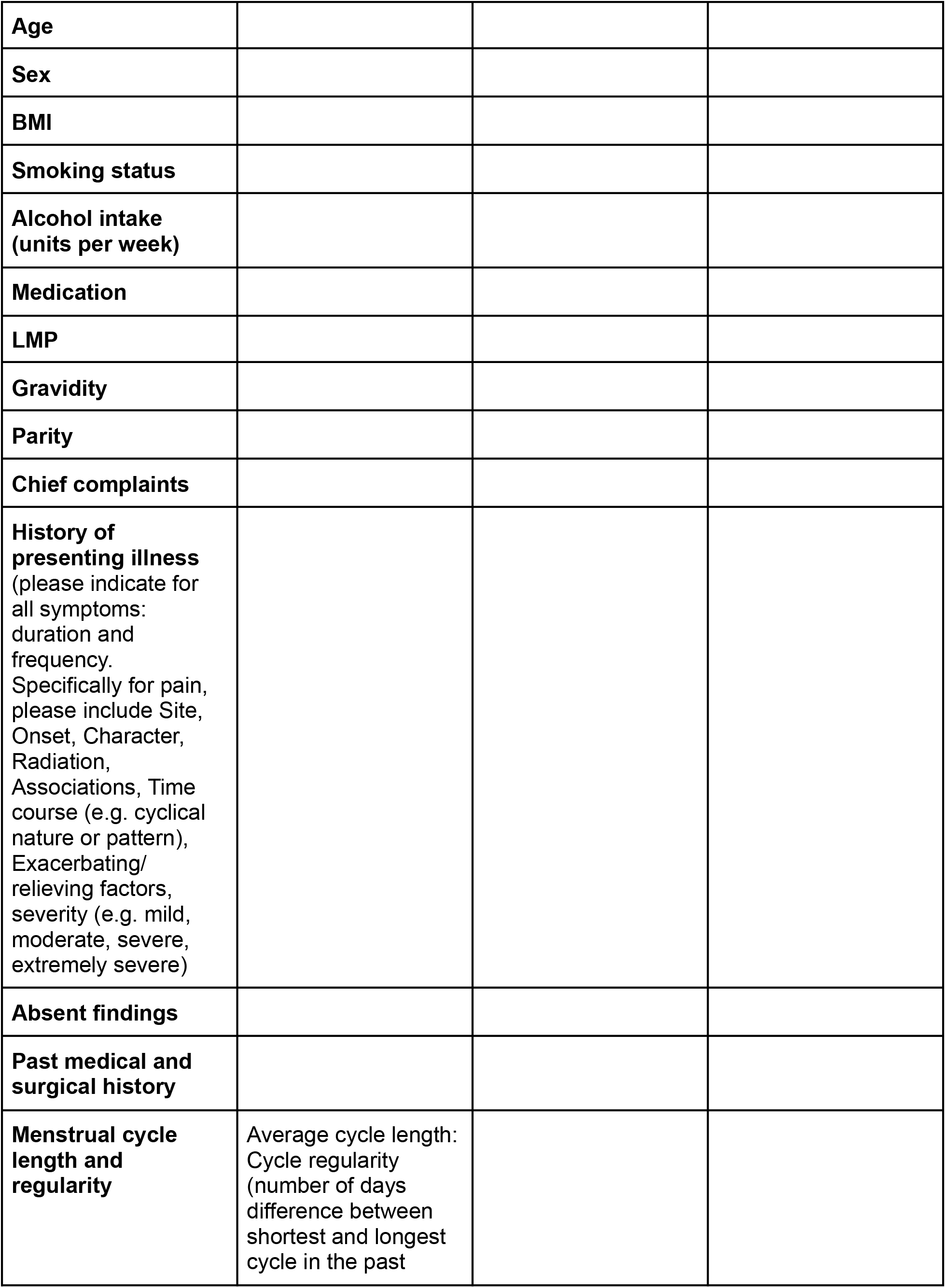
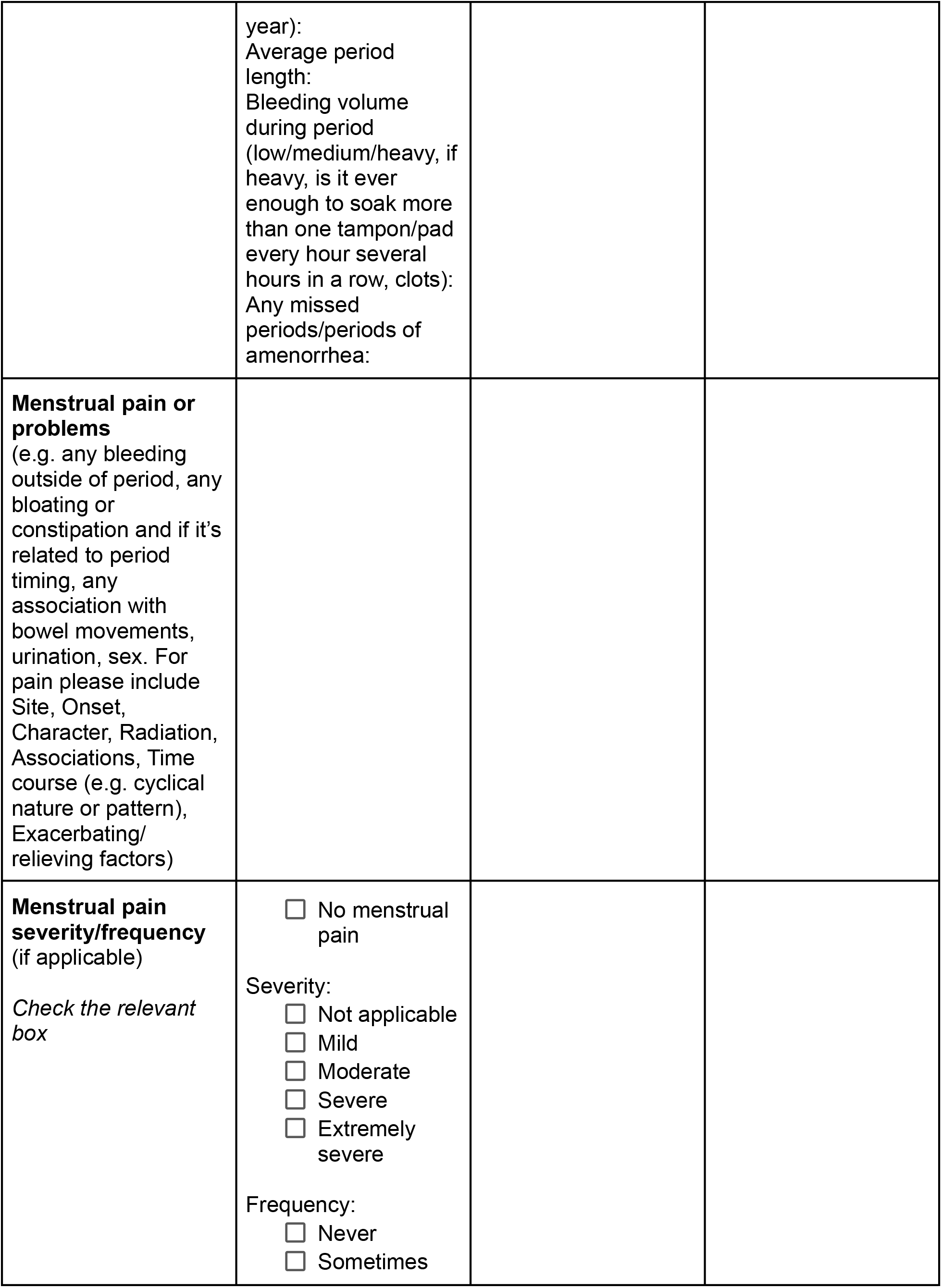
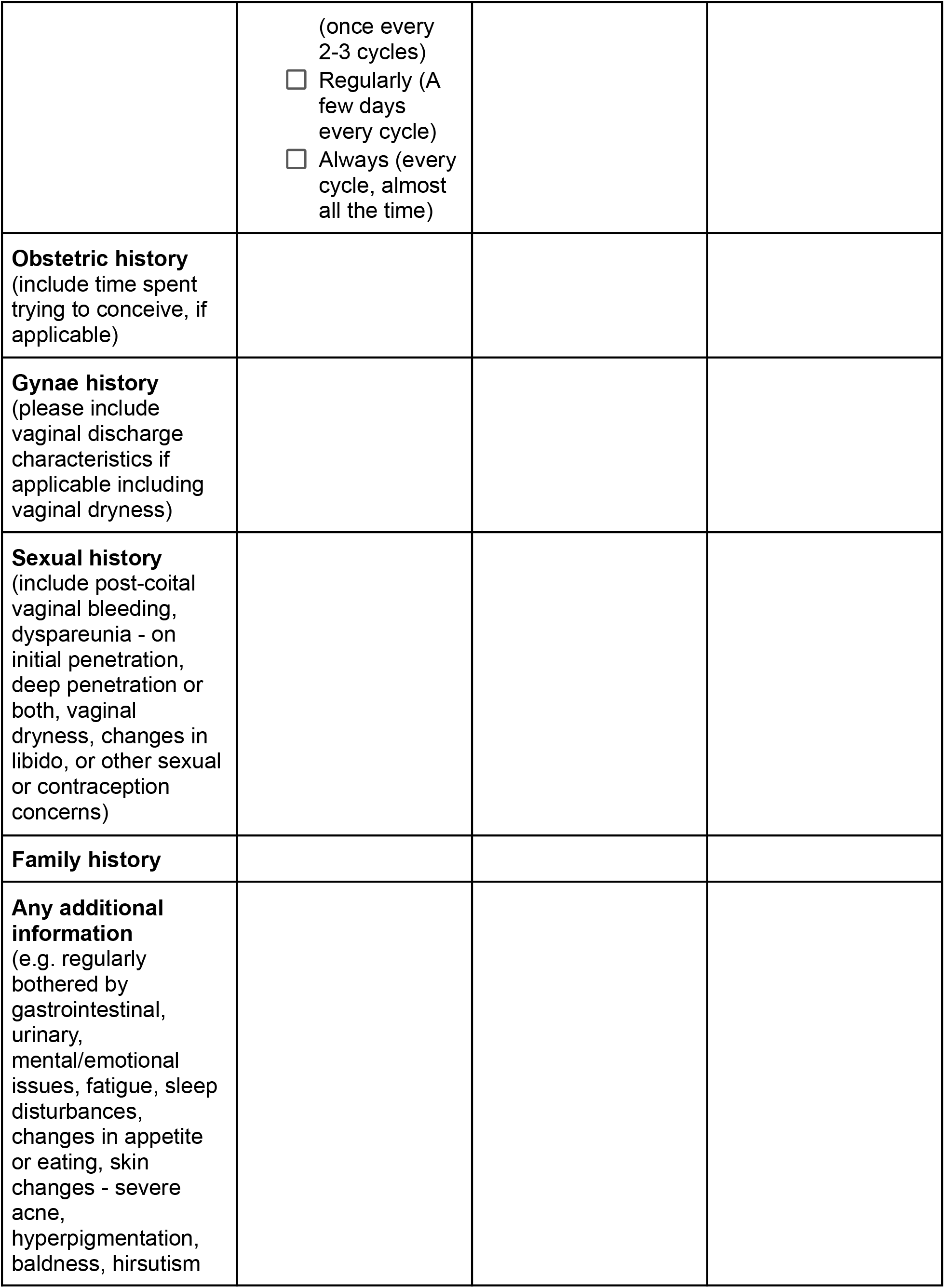
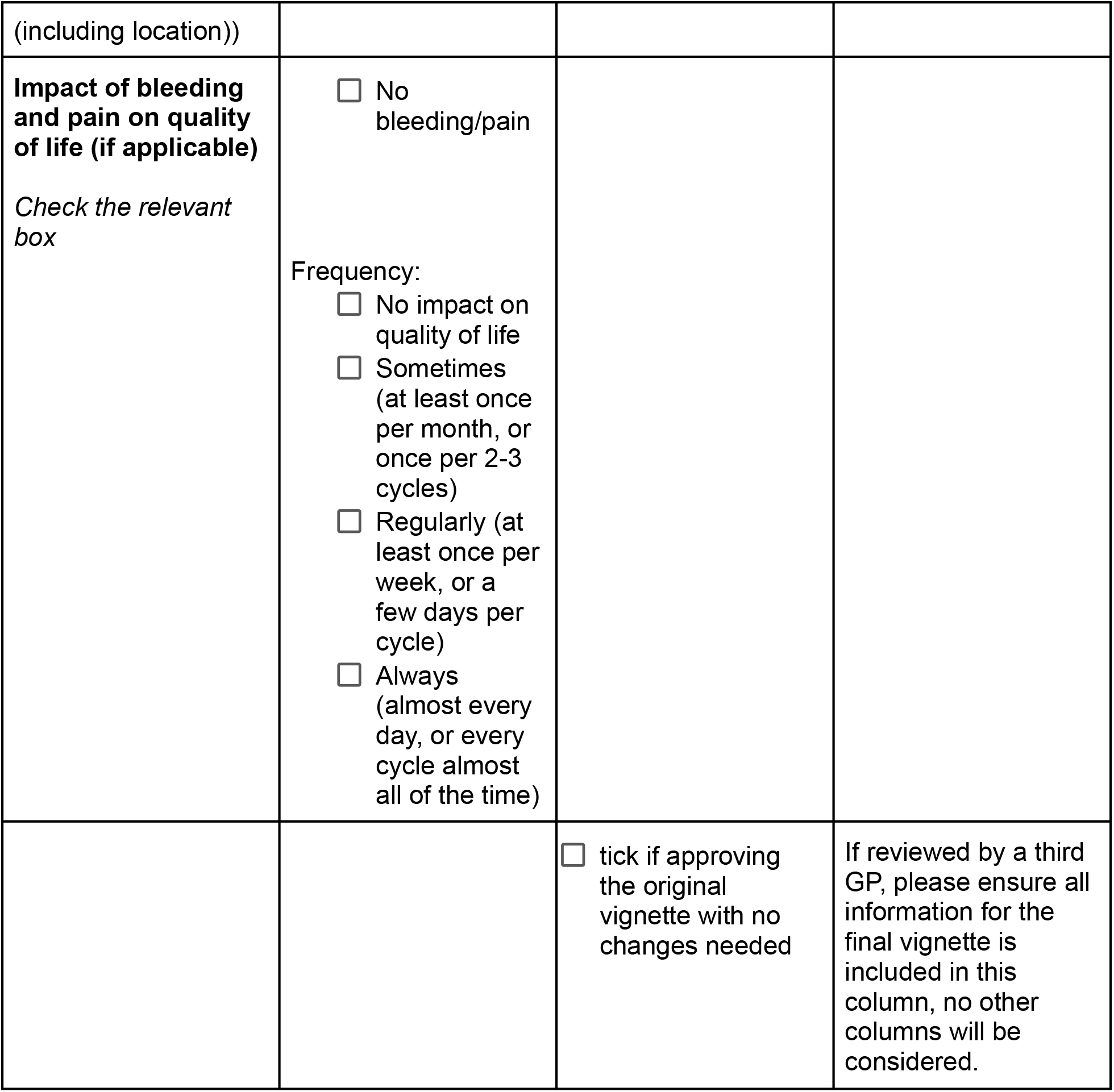
vignette template

**Appendix 2.** Definitions and Formulas for Validation Statistics^1^

| Term | Definition | Formula (variables from Table 1) |
| --- | --- | --- |
| Percent agreement (accuracy) | The proportion of cases which were correctly classified by the symptom checker as specific signs/symptoms of the condition or as limited/no signs/symptoms of the condition | $\frac{TP+TN}{TP+FN+FP+TN}$ |
| Sensitivity | The proportion of cases who truly had specific signs/symptoms of the condition that were classified as having specific signs/symptoms of the condition by the symptom checker | $\frac{TP}{TP+FN}$ |
| Specificity | The proportion of cases who truly had limited/no signs/symptoms of the condition that were classified as having limited/no signs/symptoms of the condition by the symptom checker | $\frac{TN}{FP+TN}$ |
| PPV | The probability that a case classified as having specific signs/symptoms of the condition by the symptom checker truly had specific signs/symptoms of the condition | $\frac{TP}{TP+FP}$ |
| NPV | The probability that a case was classified as having limited/no signs/symptoms of the condition by the symptom checker truly had limited/no signs/symptoms of the condition | $\frac{TN}{FN+TN}$ |

<sup>1</sup>Rothman KJ, Greenland S, Lash TL. *Modern Epidemiology*. 3rd ed., thoroughly rev. and updated. Wolters Kluwer Health/Lippincott Williams & Wilkins; 2008.

